## Supporting information document for "The GeoTox Package: Open-source software for connecting spatiotemporal exposure to individual and population-level risk"

### 1 Supporting Information

This is the Supporting Information accompanying *The GeoTox Package: Open-source software for connecting spatiotemporal exposure to individual and population-level risk*. This document contains figures S1, S2, and S3.

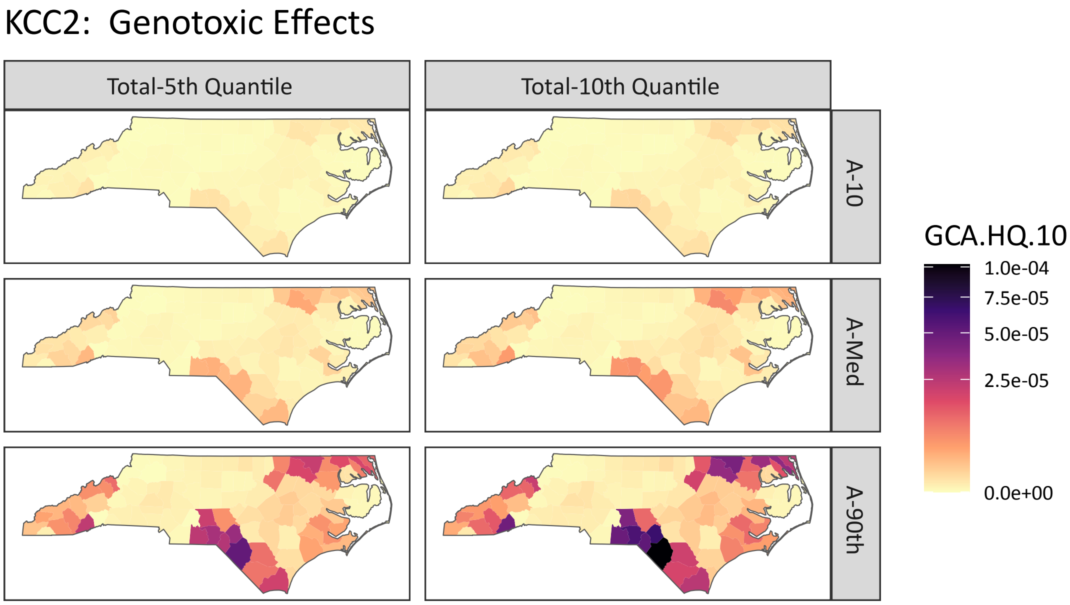


Figure S1: Multi-assay risk summaries for Key Characteristic of Carcinogens group 5 (genotoxic effects) mapped across North Carolina counties. The rows are the 10th, 50th, and 90th quantile of the assay-level hazard quotient for each county. The columns represent the second-level of summarization - the 5th and 10th quantile across all the assay-level quantiles.

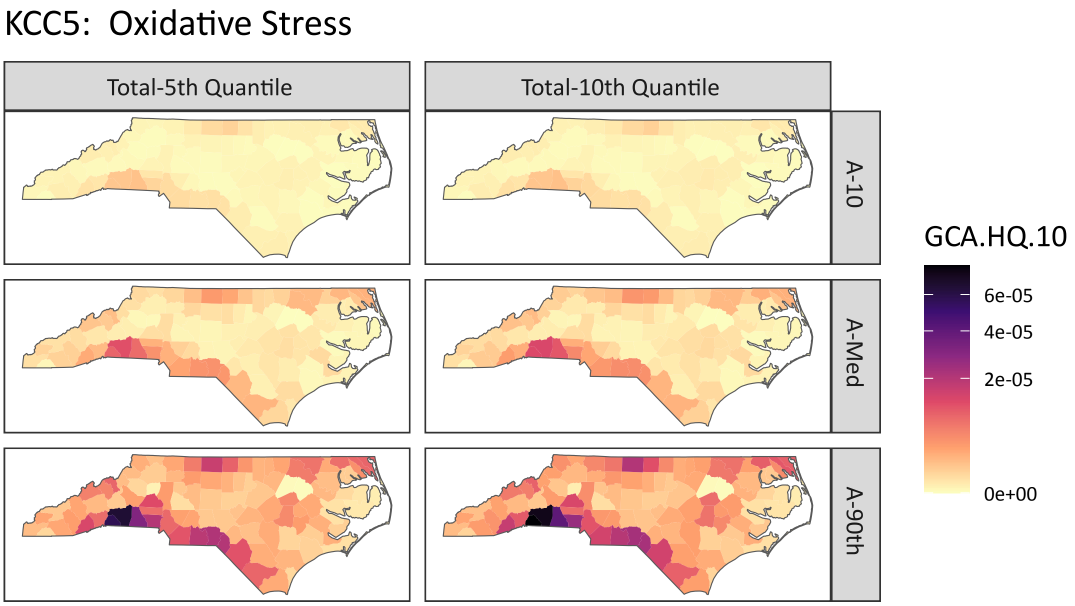


Figure S2: Multi-assay risk summaries for Key Characteristic of Carcinogens group 5 (oxidative stress) mapped across North Carolina counties. The figure panels can be interpreted in the same manner as figure S1

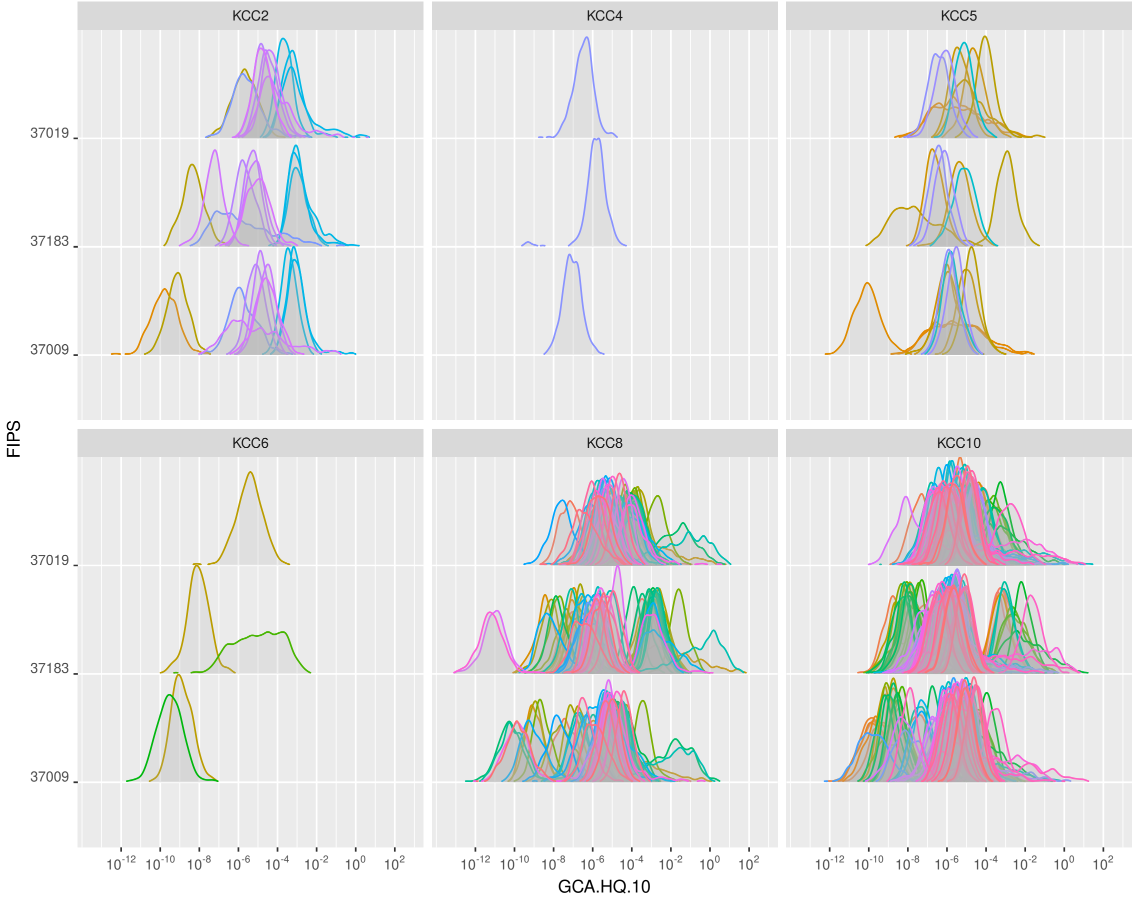


Figure S3: Hazard Quotient density with each panel as a separate KCC mode of action. Within the panels, each kernel density is the estimated hazard quotient for a given assay. The rows within the panels are the three select counties in North Carolina.
